# Prediabetic Levels of Hyperglycemia as a Predictor of Microvascular Angina with Possible Involvement of Low Levels of High-Density Lipoprotein Cholesterol and Hemoglobin

**DOI:** 10.1101/2024.08.09.24311767

**Authors:** Satoru Suzuki, Kenshi Yamanaga, Masanobu Ishii, Erika Matsumoto, Naoto Kuyama, Kyoko Hirakawa, Noriaki Tabata, Tomonori Akasaka, Koichiro Fujisue, Shinsuke Hanatani, Yuichiro Arima, Hiroki Usuku, Eiichiro Yamamoto, Kentaro Oniki, Hirofumi Soejima, Junji Saruwatari, Hiroaki Kawano, Hideaki Jinnouchi, Koichi Kaikita, Hirofumi Yasue, Hisao Ogawa, Kenichi Tsujita

## Abstract

**Background:** The management of modifiable risk factors, including conventional risk factors, can play an important role in the treatment for microvascular angina (MVA). The purpose of this study was to clarify the clinical characteristics of MVA in a case-control study, thereby elucidating the pathogenesis and management of MVA.

**Methods:** This study enrolled 92 consecutive patients with MVA (39 men, 53 women, mean age 63.2 ±12.1 years) without obstructive coronary artery disease (≥50% stenosis) or acetylcholine-provoked epicardial coronary spasms between 1993 and 2015. This study enrolled 691 age-matched Japanese participants (466 men, 225 women, mean age 62.9 ±11.0 years) without a history of chest pain or cardiovascular diseases between 2006 and 2012 as controls.

**Results:** In multivariate logistic regression analysis, elevated haemoglobin A1c (HbA1c) (per 1 mmol/mol), low high-density lipoprotein cholesterol (HDL-C) (per 1mmol/L), and low haemoglobin (Hgb) (per 10×g/L) levels were significantly associated with MVA (odds ratio [OR] 1.07[1.04-1.10], p <0.001; OR 0.13[0.06-0.29], p <0.001; OR 0.56[0.44-0.72], p <0.001, respectively). However, the difference between the prevalence of diabetes in MVA patients vs. the prevalence in controls was not significant. MVA patients without diabetes had significantly higher HbA1c levels than control patients without diabetes (39±4 mmol/mol vs. 34±3 mmol/mol, respectively; p <0.001). These results were also true for patients and controls stratified by sex. A higher prevalence of proteinuria and lower mean corpuscular volume of erythrocytes were found in MVA patients than in controls stratified by sex.

**Conclusions:** Hyperglycaemia reflected by mildly elevated HbA1c levels, (i.e., prediabetes) may be a risk factor for MVA. Low levels of HDL-C and Hgb may also be risk factors for MVA. MVA could be one of the systemic manifestations of microvascular diseases. The study may aid in identifying high-risk MVA patients, allowing increased monitoring and treatments for them.

**What is already known on this topic:** Microvascular angina (MVA) has been attracting attention of clinical research because MVA can be associated with future cardiovascular events. The management of modifiable risk factors, including conventional risk factors, can play an important role in the treatment of MVA because there is currently no specific treatment of MVA. The purpose of this study was to clarify the clinical characteristics of MVA.

**What this study adds:** Hyperglycaemia reflected by mildly elevated haemoglobin A1c levels, (i.e., prediabetes) may be a risk factor for MVA. Low levels of high-density lipoprotein cholesterol and haemoglobin may also be risk factors for MVA. A higher prevalence of proteinuria and lower mean corpuscular volume of erythrocytes were found in MVA patients than in controls stratified by sex.

**How this study might affect research, practice or policy:** MVA could be one of the systemic manifestations of microvascular diseases. The study findings may aid in identifying high-risk patients with MVA, allowing increased monitoring and treatments for them.

## Introduction

Patients with suspected angina are frequently found to have non-obstructive coronary artery disease (CAD). This condition has recently been called “ischaemia with non-obstructive CAD (INOCA)” ^1–5^. Non-acetylcholine (ACh)-provoked epicardial coronary spasm is sometimes seen in patients with INOCA. A possible cause of this can be microvascular angina (MVA) due to coronary microvascular dysfunction (CMD). MVA has been a recent focus of clinical research because MVA has emerged as an important factor associated with adverse cardiovascular outcomes ^1^ ^2^ ^6^ ^7^. However, the pathogenesis and management of MVA remain unknown.

Previous studies have found that ageing, smoking, hypertension (HT), diabetes mellitus (DM), and dyslipidaemia (DLP) could be associated with MVA ^8–10^. In contrast, other studies have found that DM was uncommon in patients with angina and non-obstructive CAD, whereas HT and DLP were relatively common ^11^ ^12^. Thus, the association between conventional cardiovascular risk factors and MVA has not yet been established ^4^. The management of modifiable risk factors including conventional risk factors can play an important role in the treatment of MVA, because there is currently no specific treatment of MVA ^4^ ^13^.

The purpose of this study was to clarify the clinical characteristics of MVA in a case-control study, thereby elucidating the pathogenesis and management of MVA.

## Methods

### Study patients and definition of MVA

We retrospectively enrolled 92 consecutive Japanese patients with MVA (39 men, 53 women) who were admitted to our institution for coronary angiography because of anginal chest pain between January 1993 and December 2015 (figure 1). All vasoactive drugs, including calcium channel blockers, nitrates, nicorandil, alpha-adrenergic blockers, and beta-adrenergic blockers were discontinued ≥ 48 hours before coronary angiography (CAG), which included an ACh provocation test and a coronary flow reserve (CFR) measurement by intravenous infusion of adenosine triphosphate (ATP). Every study patient underwent CAG in the morning while fasting. After the ACh provocation test, isosorbide dinitrate (ISDN) was infused into the left and right coronary arteries, followed by examining the epicardial coronary artery for stenosis.

**Figure 1.**
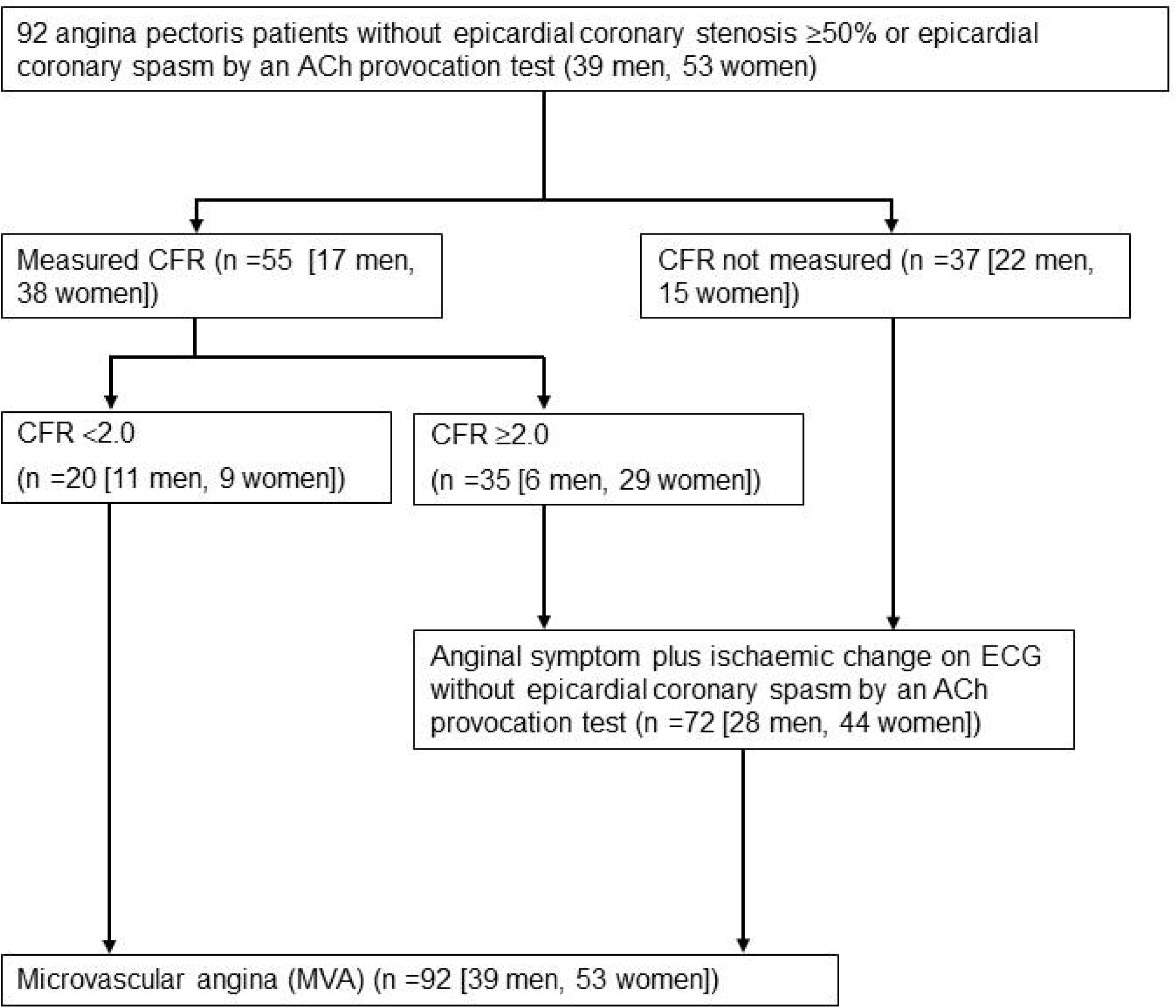
Flow chart of patient enrollment. ACh, acetylcholine; CFR, coronary flow reserve.

The diagnosis of MVA was based on satisfying all the criteria for MVA proposed by the *Coronary Vasomotion Disorders International Study Group* (COVADIS), as follows: (1) signs and/or symptoms of myocardial ischaemia, (2) absence of obstructive CAD (≥50% stenosis) by CAG, (3) objective evidence of myocardial ischaemia (i.e., chest pain and ischaemia on ECG) with non-epicardial coronary spasm by an ACh-provocation test, and (4) evidence of impaired coronary microvascular function (CFR <2.0 by invasive measurement or non-epicardial coronary spasm during an ACh-provocation test) ^14^.

Study exclusion criteria were as follows: decompensated heart failure, cardiomyopathy, severe valvular heart disease, a previous history of coronary artery revascularization, and obstructive CAD (≥50% stenosis). Patients were also excluded for a diagnosis of epicardial coronary spasm that was based on JCS/CVIT/JCC 2023 Guideline Focussed Update on Diagnosis and Treatment of Vasospastic Angina (Coronary Spastic Angina) and Coronary

Microvascular Dysfunction by the Japanese Circulation Society ^1^. In addition, patients were excluded for other factors as follows: receiving haemodialysis; or having advanced chronic obstructive pulmonary disease, advanced collagen disease, active inflammatory disease, severe liver dysfunction, and/or neoplasms at the time of enrolment.

The control patients included 691 age-matched Japanese participants (466 men, 225 women) without a history of chest pain or cardiovascular diseases who underwent medical examinations at the Japanese Red Cross Kumamoto Health Care Centre between January 2006 and April 2012.

### Measurement of coronary flow reserve

Before the ACh provocation test, we inserted a 0.014-inch guidewire that was equipped with a Doppler velocity probe (FloWire; Philips Volcano, San Diego, CA) into the left anterior descending coronary artery. After the ACh provocation test was followed by the administration of intracoronary ISDN, ATP (150 μg/kg/min) was administered via the central vein until maximal hyperaemia was achieved to allow the calculation of haemodynamic parameters. When maximal coronary hyperaemia was attained, the FloWire was used to measure the CFR, as previously described ^15^.

### Definition of risk factors for cardiovascular diseases

The conventional risk factors for cardiovascular diseases were defined as follows: current smoking, HT (blood pressure ≥140/90 mmHg or taking anti-hypertensive drugs), DLP (low-density lipoprotein cholesterol [LDL-C] ≥3.6 mmol/L [140 mg/dL], high-density lipoprotein cholesterol [HDL-C] <1.0 mmol/L [40 mg/dL], triglycerides [TG] ≥1.7 mmol/L [150 mg/dL] or taking lipid-lowering drugs), and DM (fasting plasma glucose levels ≥7.0 mmol/L [126 mg/dL], >11.1 mmol/L [200 mg/dL] during a 75-g oral glucose tolerance test, or taking anti-diabetic drugs). In regard to serum glycosylated haemoglobin A1c (HbA1c) levels, until the end of March 2012, the measurements proposed by the Japan Diabetes Society (JDS) were applied for study patients; whereas starting in April 2012, the measurements described by the National Glycohemoglobin Standardisation Programme (NGSP) were adopted. In our study, the HbA1c levels (JDS) measured until March 2012 were adjusted to the NGSP levels (i.e., HbA1c [NGSP] [%] =HbA1c [JDS] [%] +0.4) ^16^. Additionally, the HbA1c levels (NGSP) (%) were adjusted to the HbA1c levels (IFCC) (mmol/mol) (i.e., HbA1c [IFCC] [mmol/mol] = 10.93 × HbA1c [NGSP] (%) −23.52).

### Patient and public involvement

Patients and/or the public were not involved in the design, or conduct, or reporting, or dissemination plans of this research. We plan to disseminate the findings to participants and the public.

### Statistical analysis

Continuous values were expressed as means ± standard deviation (SD), whereas data with a skewed distribution were expressed as medians (interquartile range). Continuous variables were compared by two-tailed unpaired t-tests. A Mann-Whitney U test was performed for continuous variables with a skewed distribution. Frequencies of clinical variables stratified into two groups were compared by the Fisher exact test.

Clinical parameters were assessed by univariate and multivariate logistic regression analyses for significant association with MVA. The multivariate logistic regression analysis was used to evaluate parameters that were identified as statistically significant (p <0.05) by univariate logistic regression analysis, with the exclusion of redundant and/or missing data on the variables. We did not include ‘DLP’ in the multivariate logistic regression analysis because the correlation between ‘DLP’ and HDL-C levels was strong. The variables ‘current smoking’ and ‘proteinuria’ were not included in the multivariate logistic regression analysis because there were only a few patients who were current smokers or had proteinuria (tables 3-5).

Stata version 18 (Stata Corp., College Station, TX, USA) and BellCurve for Excel, version 4.06 (Social Survey Research Information Co., Ltd., Tokyo, Japan) were used for statistical analysis. Statistical significance was defined as a p-value < 0.05.

## Results

### Clinical characteristics

Table 1 shows the clinical characteristics of 92 (39 men, 53 women) patients with MVA and 691 (466 men, 225 women) control participants. The patients with MVA had a significantly lower prevalence of DLP; higher HbA1c, lower HDL-C, higher CRP, and lower Hgb levels; and a higher prevalence of proteinuria than the control participants.

**Table 1.**
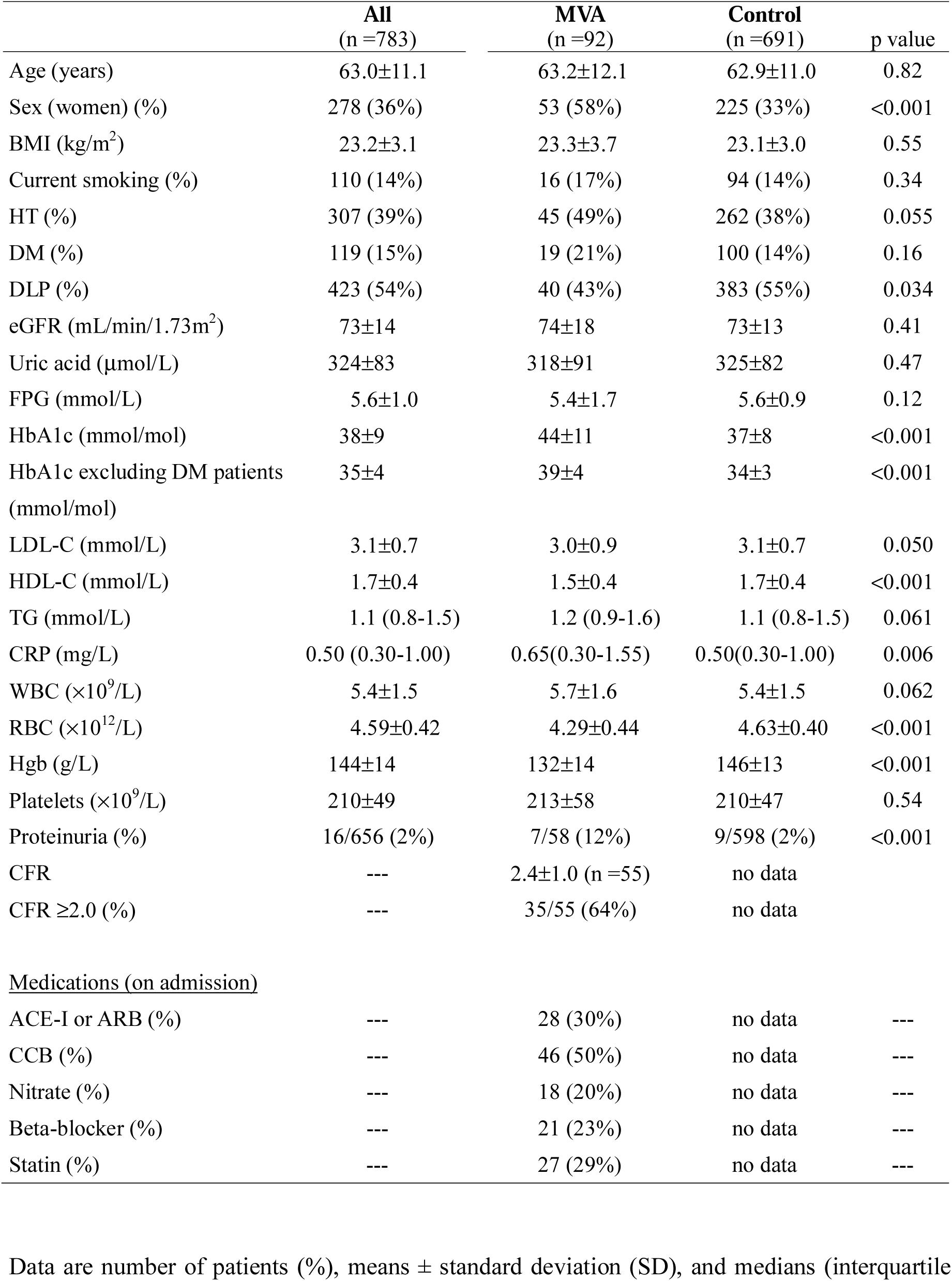
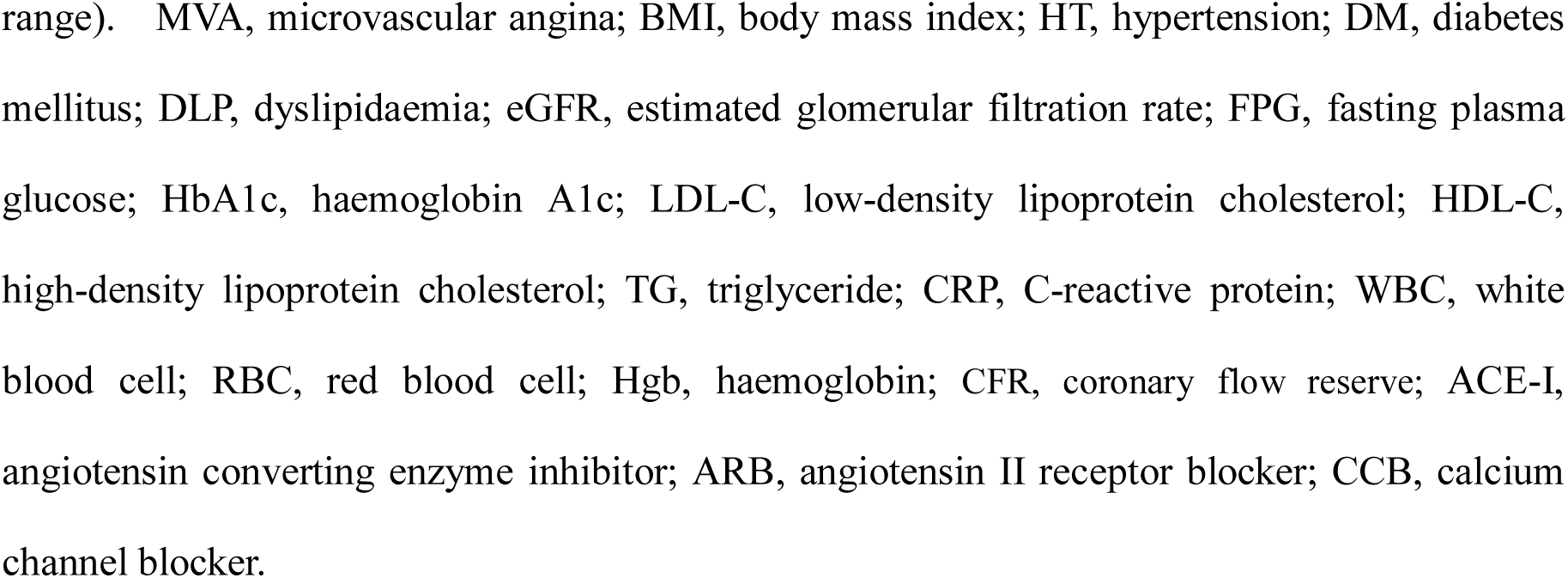
Clinical Characteristics of MVA Patients and Control Participants.

|  | **All**<br>(n =783) | **MVA**<br>(n =92) | **Control**<br>(n =691) | p value |
| --- | --- | --- | --- | --- |
| Age (years) | 63.0±11.1 | 63.2±12.1 | 62.9±11.0 | 0.82 |
| Sex (women) (%) | 278 (36%) | 53 (58%) | 225 (33%) | <0.001 |
| BMI (kg/m<sup>2</sup>) | 23.2±3.1 | 23.3±3.7 | 23.1±3.0 | 0.55 |
| Current smoking (%) | 110 (14%) | 16 (17%) | 94 (14%) | 0.34 |
| HT (%) | 307 (39%) | 45 (49%) | 262 (38%) | 0.055 |
| DM (%) | 119 (15%) | 19 (21%) | 100 (14%) | 0.16 |
| DLP (%) | 423 (54%) | 40 (43%) | 383 (55%) | 0.034 |
| eGFR (mL/min/1.73m<sup>2</sup>) | 73±14 | 74±18 | 73±13 | 0.41 |
| Uric acid (μmol/L) | 324±83 | 318±91 | 325±82 | 0.47 |
| FPG (mmol/L) | 5.6±1.0 | 5.4±1.7 | 5.6±0.9 | 0.12 |
| HbA1c (mmol/mol) | 38±9 | 44±11 | 37±8 | <0.001 |
| HbA1c excluding DM patients<br>(mmol/mol) | 35±4 | 39±4 | 34±3 | <0.001 |
| LDL-C (mmol/L) | 3.1±0.7 | 3.0±0.9 | 3.1±0.7 | 0.050 |
| HDL-C (mmol/L) | 1.7±0.4 | 1.5±0.4 | 1.7±0.4 | <0.001 |
| TG (mmol/L) | 1.1 (0.8-1.5) | 1.2 (0.9-1.6) | 1.1 (0.8-1.5) | 0.061 |
| CRP (mg/L) | 0.50 (0.30-1.00) | 0.65(0.30-1.55) | 0.50(0.30-1.00) | 0.006 |
| WBC (×10<sup>9</sup>/L) | 5.4±1.5 | 5.7±1.6 | 5.4±1.5 | 0.062 |
| RBC (×10<sup>12</sup>/L) | 4.59±0.42 | 4.29±0.44 | 4.63±0.40 | <0.001 |
| Hgb (g/L) | 144±14 | 132±14 | 146±13 | <0.001 |
| Platelets (×10<sup>9</sup>/L) | 210±49 | 213±58 | 210±47 | 0.54 |
| Proteinuria (%) | 16/656 (2%) | 7/58 (12%) | 9/598 (2%) | <0.001 |
| CFR | --- | 2.4±1.0 (n =55) | no data |  |
| CFR ≥2.0 (%) | --- | 35/55 (64%) | no data |  |
| **<u>Medications (on admission)</u>** |  |  |  |  |
| ACE-I or ARB (%) | --- | 28 (30%) | no data | --- |
| CCB (%) | --- | 46 (50%) | no data | --- |
| Nitrate (%) | --- | 18 (20%) | no data | --- |
| Beta-blocker (%) | --- | 21 (23%) | no data | --- |
| Statin (%) | --- | 27 (29%) | no data | --- |

Data are number of patients (%), means ± standard deviation (SD), and medians (interquartile
range). MVA, microvascular angina; BMI, body mass index; HT, hypertension; DM, diabetes mellitus; DLP, dyslipidaemia; eGFR, estimated glomerular filtration rate; FPG, fasting plasma glucose; HbA1c, haemoglobin A1c; LDL-C, low-density lipoprotein cholesterol; HDL-C, high-density lipoprotein cholesterol; TG, triglyceride; CRP, C-reactive protein; WBC, white blood cell; RBC, red blood cell; Hgb, haemoglobin; CFR, coronary flow reserve; ACE-I, angiotensin converting enzyme inhibitor; ARB, angiotensin II receptor blocker; CCB, calcium channel blocker.

Table 2 shows the clinical characteristics of MVA patients and control participants stratified by sex. The men patients with MVA had significantly higher HbA1c, lower HDL-C, and lower Hgb levels; and a higher prevalence of proteinuria than the control participants. The women patients with MVA had significantly higher HbA1c, lower HDL-C, higher TG, higher CRP, higher WBC, and lower Hgb levels; and higher prevalence of current smoking, and higher prevalence of proteinuria than the control participants.

**Table 2.**
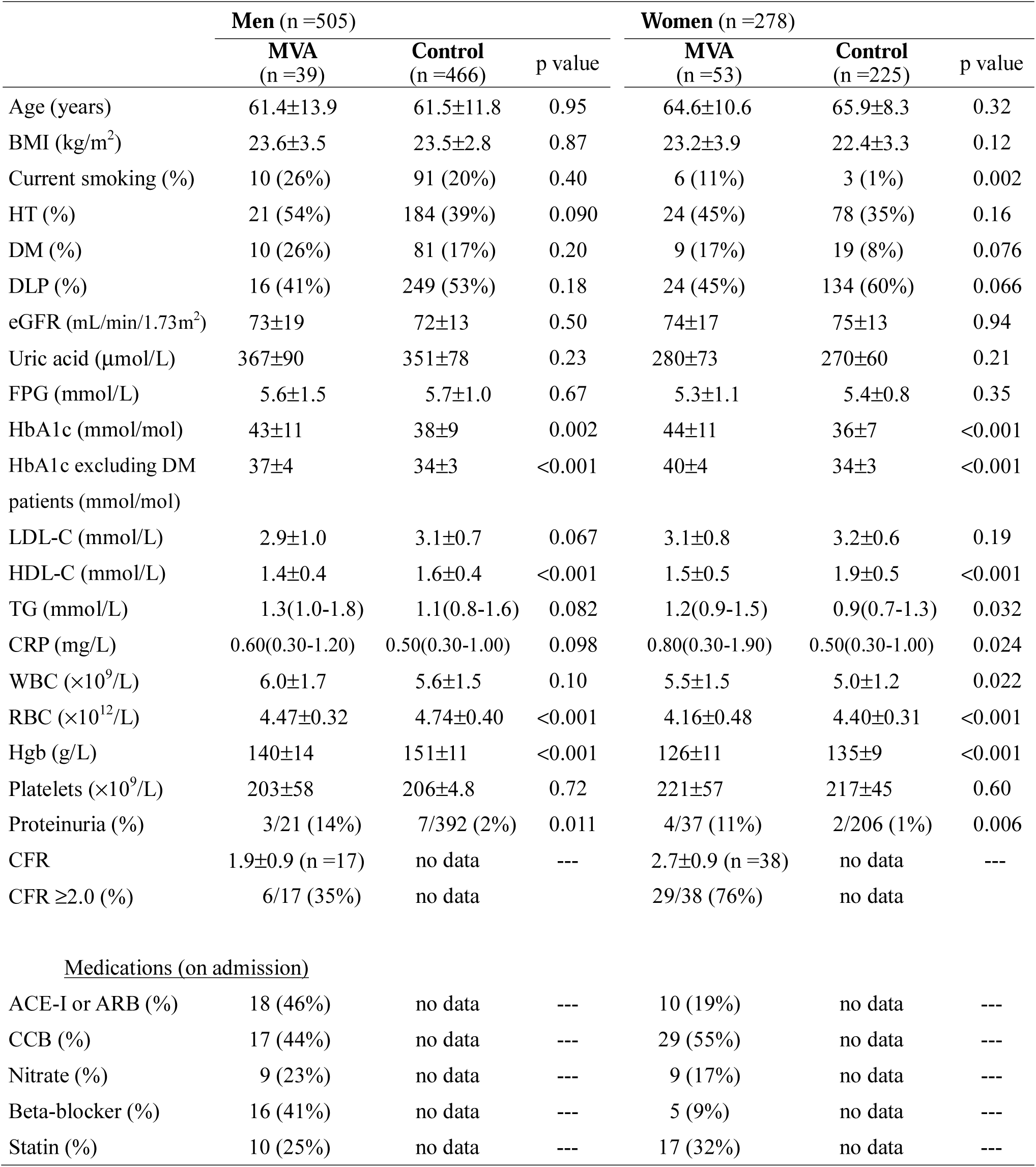
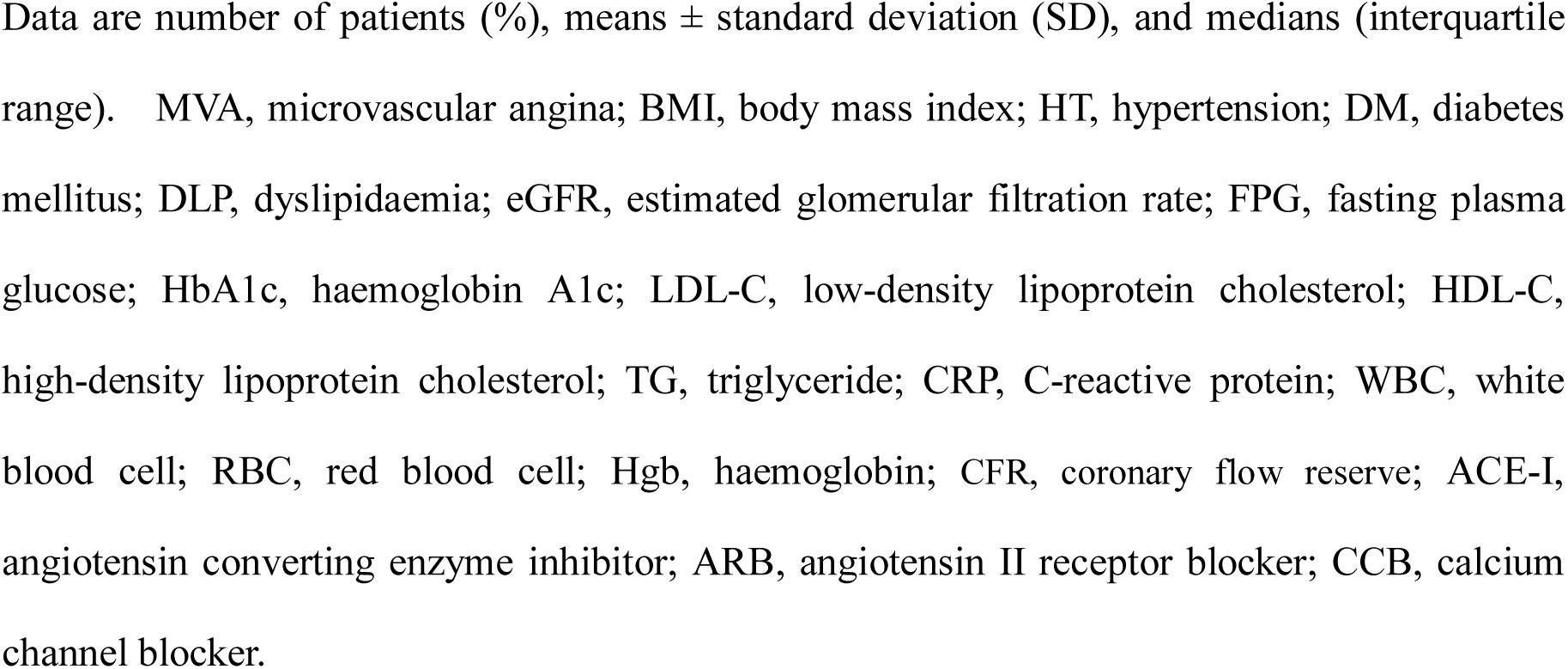
Clinical Characteristics of MVA Patients and Control Participants in Men and Women.

|  | Men (n =505) |  |  | Women (n =278) |  |  |
| --- | --- | --- | --- | --- | --- | --- |
|  | MVA<br>(n =39) | Control<br>(n =466) | p value | MVA<br>(n =53) | Control<br>(n =225) | p value |
| Age (years) | 61.4±13.9 | 61.5±11.8 | 0.95 | 64.6±10.6 | 65.9±8.3 | 0.32 |
| BMI (kg/m<sup>2</sup>) | 23.6±3.5 | 23.5±2.8 | 0.87 | 23.2±3.9 | 22.4±3.3 | 0.12 |
| Current smoking (%) | 10 (26%) | 91 (20%) | 0.40 | 6 (11%) | 3 (1%) | 0.002 |
| HT (%) | 21 (54%) | 184 (39%) | 0.090 | 24 (45%) | 78 (35%) | 0.16 |
| DM (%) | 10 (26%) | 81 (17%) | 0.20 | 9 (17%) | 19 (8%) | 0.076 |
| DLP (%) | 16 (41%) | 249 (53%) | 0.18 | 24 (45%) | 134 (60%) | 0.066 |
| eGFR (mL/min/1.73m<sup>2</sup>) | 73±19 | 72±13 | 0.50 | 74±17 | 75±13 | 0.94 |
| Uric acid (μmol/L) | 367±90 | 351±78 | 0.23 | 280±73 | 270±60 | 0.21 |
| FPG (mmol/L) | 5.6±1.5 | 5.7±1.0 | 0.67 | 5.3±1.1 | 5.4±0.8 | 0.35 |
| HbA1c (mmol/mol) | 43±11 | 38±9 | 0.002 | 44±11 | 36±7 | <0.001 |
| HbA1c excluding DM patients (mmol/mol) | 37±4 | 34±3 | <0.001 | 40±4 | 34±3 | <0.001 |
| LDL-C (mmol/L) | 2.9±1.0 | 3.1±0.7 | 0.067 | 3.1±0.8 | 3.2±0.6 | 0.19 |
| HDL-C (mmol/L) | 1.4±0.4 | 1.6±0.4 | <0.001 | 1.5±0.5 | 1.9±0.5 | <0.001 |
| TG (mmol/L) | 1.3(1.0-1.8) | 1.1(0.8-1.6) | 0.082 | 1.2(0.9-1.5) | 0.9(0.7-1.3) | 0.032 |
| CRP (mg/L) | 0.60(0.30-1.20) | 0.50(0.30-1.00) | 0.098 | 0.80(0.30-1.90) | 0.50(0.30-1.00) | 0.024 |
| WBC (×10<sup>9</sup>/L) | 6.0±1.7 | 5.6±1.5 | 0.10 | 5.5±1.5 | 5.0±1.2 | 0.022 |
| RBC (×10<sup>12</sup>/L) | 4.47±0.32 | 4.74±0.40 | <0.001 | 4.16±0.48 | 4.40±0.31 | <0.001 |
| Hgb (g/L) | 140±14 | 151±11 | <0.001 | 126±11 | 135±9 | <0.001 |
| Platelets (×10<sup>9</sup>/L) | 203±58 | 206±4.8 | 0.72 | 221±57 | 217±45 | 0.60 |
| Proteinuria (%) | 3/21 (14%) | 7/392 (2%) | 0.011 | 4/37 (11%) | 2/206 (1%) | 0.006 |
| CFR | 1.9±0.9 (n =17) | no data | --- | 2.7±0.9 (n =38) | no data | --- |
| CFR ≥2.0 (%) | 6/17 (35%) | no data |  | 29/38 (76%) | no data |  |
| **Medications (on admission)** |  |  |  |  |  |  |
| ACE-I or ARB (%) | 18 (46%) | no data | --- | 10 (19%) | no data | --- |
| CCB (%) | 17 (44%) | no data | --- | 29 (55%) | no data | --- |
| Nitrate (%) | 9 (23%) | no data | --- | 9 (17%) | no data | --- |
| Beta-blocker (%) | 16 (41%) | no data | --- | 5 (9%) | no data | --- |
| Statin (%) | 10 (25%) | no data | --- | 17 (32%) | no data | --- |

Data are number of patients (%), means ± standard deviation (SD), and medians (interquartile range). MVA, microvascular angina; BMI, body mass index; HT, hypertension; DM, diabetes mellitus; DLP, dyslipidaemia; eGFR, estimated glomerular filtration rate; FPG, fasting plasma glucose; HbA1c, haemoglobin A1c; LDL-C, low-density lipoprotein cholesterol; HDL-C, high-density lipoprotein cholesterol; TG, triglyceride; CRP, C-reactive protein; WBC, white blood cell; RBC, red blood cell; Hgb, haemoglobin; CFR, coronary flow reserve; ACE-I, angiotensin converting enzyme inhibitor; ARB, angiotensin II receptor blocker; CCB, calcium channel blocker.

### Univariate and multivariate logistic regression analysis of characteristics associated with MVA

Table 3 shows the results of univariate and multivariate logistic regression analysis for the association of variables with MVA. Univariate logistic regression analysis showed that women, HT, DLP, HbA1c, HDL-C, Hgb, and proteinuria were all significantly associated with MVA. Multivariate logistic regression analysis showed that HbA1c, HDL-C, and Hgb were significantly associated with MVA.

**Table 3.**
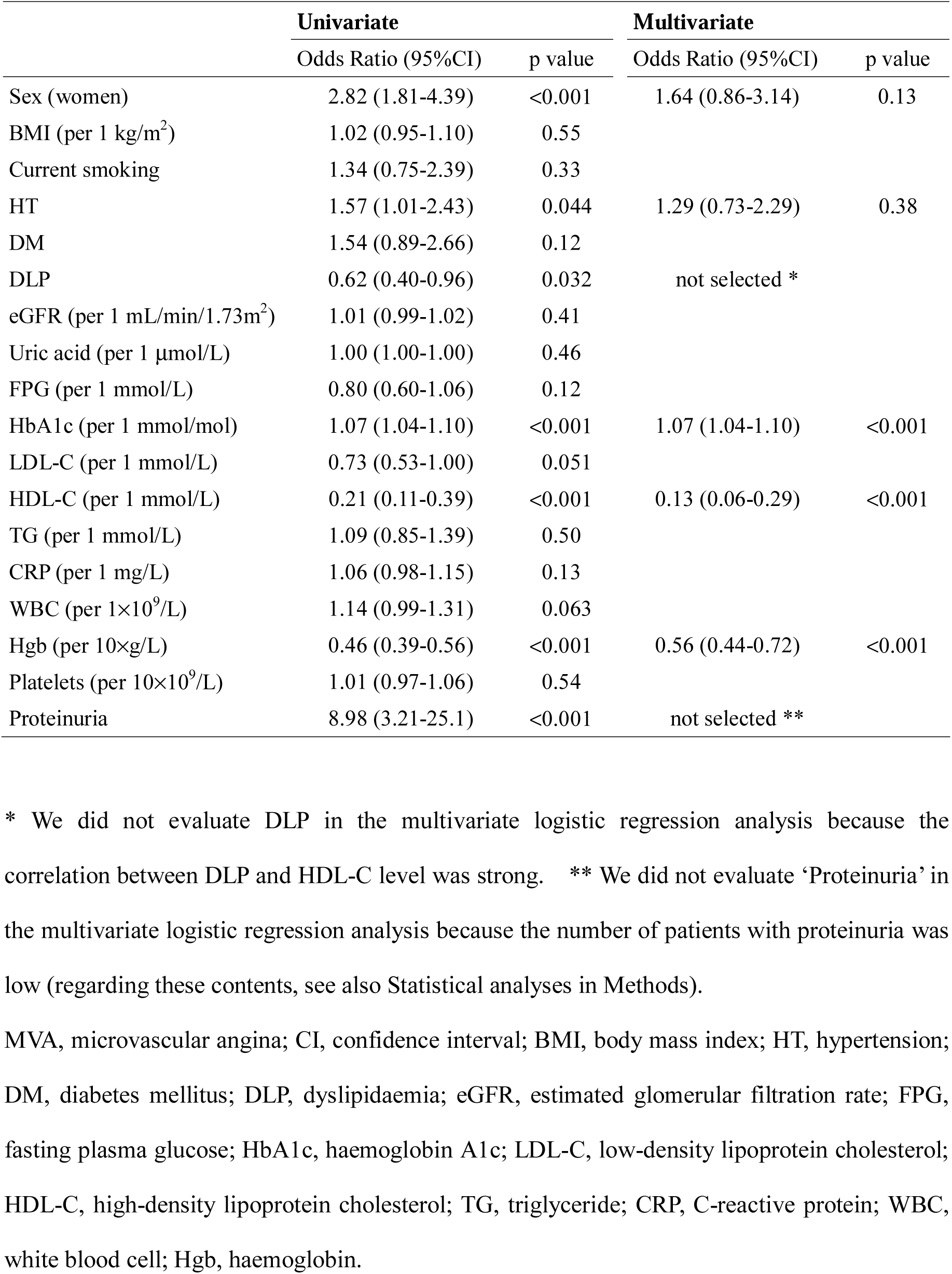
Univariate and Multivariate Logistic Regression Analysis for MVA.

|  | **Univariate** |  | **Multivariate** |  |
| --- | --- | --- | --- | --- |
|  | Odds Ratio (95%CI) | p value | Odds Ratio (95%CI) | p value |
| Sex (women) | 2.82 (1.81-4.39) | <0.001 | 1.64 (0.86-3.14) | 0.13 |
| BMI (per 1 kg/m<sup>2</sup>) | 1.02 (0.95-1.10) | 0.55 |  |  |
| Current smoking | 1.34 (0.75-2.39) | 0.33 |  |  |
| HT | 1.57 (1.01-2.43) | 0.044 | 1.29 (0.73-2.29) | 0.38 |
| DM | 1.54 (0.89-2.66) | 0.12 |  |  |
| DLP | 0.62 (0.40-0.96) | 0.032 | not selected * |  |
| eGFR (per 1 mL/min/1.73m<sup>2</sup>) | 1.01 (0.99-1.02) | 0.41 |  |  |
| Uric acid (per 1 µmol/L) | 1.00 (1.00-1.00) | 0.46 |  |  |
| FPG (per 1 mmol/L) | 0.80 (0.60-1.06) | 0.12 |  |  |
| HbA1c (per 1 mmol/mol) | 1.07 (1.04-1.10) | <0.001 | 1.07 (1.04-1.10) | <0.001 |
| LDL-C (per 1 mmol/L) | 0.73 (0.53-1.00) | 0.051 |  |  |
| HDL-C (per 1 mmol/L) | 0.21 (0.11-0.39) | <0.001 | 0.13 (0.06-0.29) | <0.001 |
| TG (per 1 mmol/L) | 1.09 (0.85-1.39) | 0.50 |  |  |
| CRP (per 1 mg/L) | 1.06 (0.98-1.15) | 0.13 |  |  |
| WBC (per 1×10<sup>9</sup>/L) | 1.14 (0.99-1.31) | 0.063 |  |  |
| Hgb (per 10×g/L) | 0.46 (0.39-0.56) | <0.001 | 0.56 (0.44-0.72) | <0.001 |
| Platelets (per 10×10<sup>9</sup>/L) | 1.01 (0.97-1.06) | 0.54 |  |  |
| Proteinuria | 8.98 (3.21-25.1) | <0.001 | not selected ** |  |

\* We did not evaluate DLP in the multivariate logistic regression analysis because the correlation between DLP and HDL-C level was strong. \*\* We did not evaluate 'Proteinuria' in the multivariate logistic regression analysis because the number of patients with proteinuria was low (regarding these contents, see also Statistical analyses in Methods).
MVA, microvascular angina; CI, confidence interval; BMI, body mass index; HT, hypertension; DM, diabetes mellitus; DLP, dyslipidaemia; eGFR, estimated glomerular filtration rate; FPG, fasting plasma glucose; HbA1c, haemoglobin A1c; LDL-C, low-density lipoprotein cholesterol; HDL-C, high-density lipoprotein cholesterol; TG, triglyceride; CRP, C-reactive protein; WBC, white blood cell; Hgb, haemoglobin.

Table 4 shows the results of univariate and multivariate logistic regression analysis for MVA in men. Univariate logistic regression analysis showed that HbA1c, HDL-C, Hgb, and proteinuria were all significantly associated with MVA. Multivariate logistic regression analysis showed that HbA1c, HDL-C, and Hgb were significantly associated with MVA.

**Table 4.** Univariate and Multivariate Logistic Regression Analysis for MVA in Men.

|  | Univariate |  | Multivariate |  |
| --- | --- | --- | --- | --- |
|  | Odds Ratio (95%CI) | p value | Odds Ratio (95%CI) | p value |
| BMI (per 1 kg/m<sup>2</sup>) | 1.01 (0.90-1.14) | 0.87 |  |  |
| Current smoking | 1.42 (0.67-3.02) | 0.36 |  |  |
| HT | 1.79 (0.93-3.45) | 0.083 |  |  |
| DM | 1.64 (0.77-3.50) | 0.20 |  |  |
| DLP | 0.61 (0.31-1.18) | 0.14 |  |  |
| eGFR (per 1 mL/min/1.73m<sup>2</sup>) | 1.01 (0.99-1.03) | 0.50 |  |  |
| Uric acid (per 1 µmol/L) | 1.00 (1.00-1.00) | 0.23 |  |  |
| FPG (per 1 mmol/L) | 0.93 (0.65-1.32) | 0.67 |  |  |
| HbA1c (per 1 mmol/mol) | 1.05 (1.02-1.08) | 0.004 | 1.04 (1.00-1.08) | 0.034 |
| LDL-C (per 1 mmol/L) | 0.65 (0.41-1.03) | 0.068 |  |  |
| HDL-C (per 1 mmol/L) | 0.11 (0.04-0.32) | <0.001 | 0.10 (0.03-0.34) | <0.001 |
| TG (per 1 mmol/L) | 1.28 (0.92-1.78) | 0.15 |  |  |
| CRP (per 1 mg/L) | 1.05 (0.95-1.15) | 0.34 |  |  |
| WBC (per 1×10<sup>9</sup>/L) | 1.18 (0.97-1.42) | 0.10 |  |  |
| Hgb (per 10×g/L) | 0.51 (0.39-0.66) | <0.001 | 0.64 (0.48-0.87) | 0.004 |
| Platelets (per 10×10<sup>9</sup>/L) | 1.00 (0.92-1.06) | 0.72 |  |  |
| Proteinuria | 9.17 (2.19-34.4) | 0.002 | not selected * |  |

\* We did not evaluate ‘Proteinuria’ in the multivariate logistic regression analysis because the number of patients with proteinuria was low (regarding the content, see also Statistical analyses in Methods).
MVA, microvascular angina; CI, confidence interval; BMI, body mass index; HT, hypertension; DM, diabetes mellitus; DLP, dyslipidaemia; eGFR, estimated glomerular filtration rate; FPG, fasting plasma glucose; HbA1c, haemoglobin A1c; LDL-C, low-density lipoprotein cholesterol; HDL-C, high-density lipoprotein cholesterol; TG, triglyceride; CRP, C-reactive protein; WBC, white blood cell; Hgb, haemoglobin.

Table 5 shows the results of univariate and multivariate logistic regression analysis for MVA in women. Univariate logistic regression analysis showed that current smoking, HbA1c, HDL-C, CRP, WBC, Hgb, and proteinuria were all significantly associated with MVA. Multivariate logistic regression analysis showed that HbA1c, HDL-C, WBC, and Hgb were significantly associated with MVA.

**Table 5.** Univariate and Multivariate Logistic Regression Analysis for MVA in Women.

|  | Univariate |  | Multivariate |  |
| --- | --- | --- | --- | --- |
|  | Odds Ratio (95%CI) | p value | Odds Ratio (95%CI) | p value |
| BMI (per 1 kg/m<sup>2</sup>) | 1.07 (0.98-1.16) | 0.12 |  |  |
| Current smoking | 9.45 (2.28-39.1) | 0.002 | not selected * |  |
| HT | 1.56 (0.85-2.86) | 0.15 |  |  |
| DM | 2.22 (0.94-5.23) | 0.069 |  |  |
| DLP | 0.56 (0.31-1.03) | 0.061 |  |  |
| eGFR (per 1 mL/min/1.73m<sup>2</sup>) | 1.00 (0.98-1.02) | 0.94 |  |  |
| Uric acid (per 1 µmol/L) | 1.00 (1.00-1.01) | 0.21 |  |  |
| FPG (per 1 mmol/L) | 0.87 (0.53-1.25) | 0.35 |  |  |
| HbA1c (per 1 mmol/mol) | 1.12 (1.07-1.18) | <0.001 | 1.09 (1.04-1.15) | 0.001 |
| LDL-C (per 1 mmol/L) | 0.74 (0.46-1.17) | 0.19 |  |  |
| HDL-C (per 1 mmol/L) | 0.12 (0.05-0.29) | <0.001 | 0.65 (0.49-0.86) | 0.003 |
| TG (per 1 mmol/L) | 1.09 (0.77-1.55) | 0.63 |  |  |
| CRP (per 1 mg/L) | 1.23 (1.01-1.50) | 0.042 | 0.25 (0.01-10.3) | 0.47 |
| WBC (per 1×10<sup>9</sup>/L) | 1.30 (1.04-1.62) | 0.024 | 1.50 (1.03-2.20) | 0.037 |
| Hgb (per 10×g/L) | 0.39 (0.28-0.55) | <0.001 | 0.41 (0.25-0.65) | <0.001 |
| Platelets (per 10×10<sup>9</sup>/L) | 1.02 (0.96-1.08) | 0.60 |  |  |
| Proteinuria | 12.4 (2.18-70.2) | 0.005 | not selected * |  |

\* We did not evaluate ‘current smoking’ and ‘proteinuria’ in the multivariate logistic regression analysis because the numbers of patients with current smoking or proteinuria were low (regarding these contents, see also Statistical analyses in Methods).
MVA, microvascular angina; CI, confidence interval; BMI, body mass index; HT, hypertension; DM, diabetes mellitus; DLP, dyslipidaemia; eGFR, estimated glomerular filtration rate; FPG, fasting plasma glucose; HbA1c, haemoglobin A1c; LDL-C, low-density lipoprotein cholesterol; HDL-C, high-density lipoprotein cholesterol; TG, triglyceride; CRP, C-reactive protein; WBC, white blood cell; Hgb, haemoglobin.

### Erythrocyte indices of MVA in men and women

Table 6 compares the erythrocyte indices between patients with MVA and control participants stratified by sex. Men with MVA had a significantly lower Hgb level and lower mean corpuscular volume (MCV) than the control men. Women with MVA had a significantly lower Hgb level, lower MCV, and a lower serum iron level than the control women.

**Table 6.** Erythrocyte Indices of MVA in Men and Women.

|  | **Men (n =505)** |  |  | **Women (n =278)** |  |  |
| --- | --- | --- | --- | --- | --- | --- |
|  | **MVA**<br>**(n =39)** | **Control**<br>**(n =466)** | **p value** | **MVA**<br>**(n =53)** | **Control**<br>**(n =225)** | **p value** |
| RBC ($\times 10^{12}/L$) | 4.47±0.32 | 4.74±0.40 | <0.001 | 4.16±0.48 | 4.40±0.31 | <0.001 |
| Haematocrit (L/L) | 0.41±0.04 | 0.45±0.03 | <0.001 | 0.38±0.03 | 0.41±0.03 | <0.001 |
| Hgb (g/L) | 140±14 | 151±11 | <0.001 | 126±11 | 135±9 | <0.001 |
| MCV (fL) | 91.8±4.0 | 95.1±3.9 | <0.001 | 92.0±4.0 | 93.8±3.9 | <0.001 |
| Serum iron (μmol/L) | 20±12 | 21±7 | 0.25 | 16±6 | 20±6 | <0.001 |
| Ferritin (μg/L) | 168±121 | 139±108 | 0.26 | 83±80 | 75±55 | 0.62 |

Data are number of patients (%), means ± standard deviation (SD). MVA, microvascular angina; Hgb, haemoglobin; MCV, mean corpuscular volume.

### Clinical characteristics of patients with MVA stratified by CFR <2.0 and CFR ≥2.0

Table 7 shows the clinical characteristics of CFR <2.0 and CFR ≥2.0 in 55 MVA patients whose CFRs were measured amongst 92 MVA patients. CFR ≥2.0 was more common in women with MVA than in men with MVA (29/35 [83%] vs. 9/20 [45%], p =0.006).

**Table 7.**
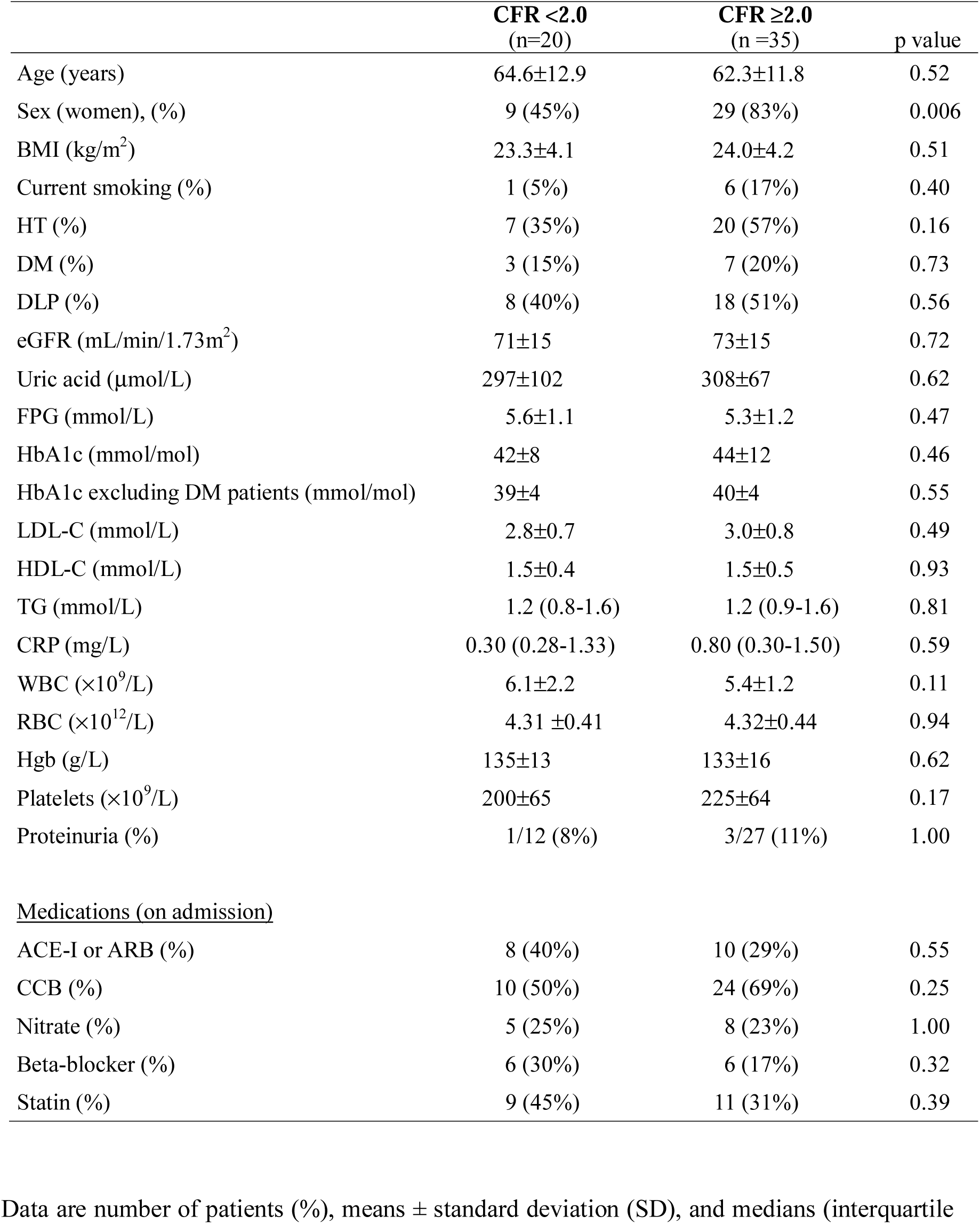
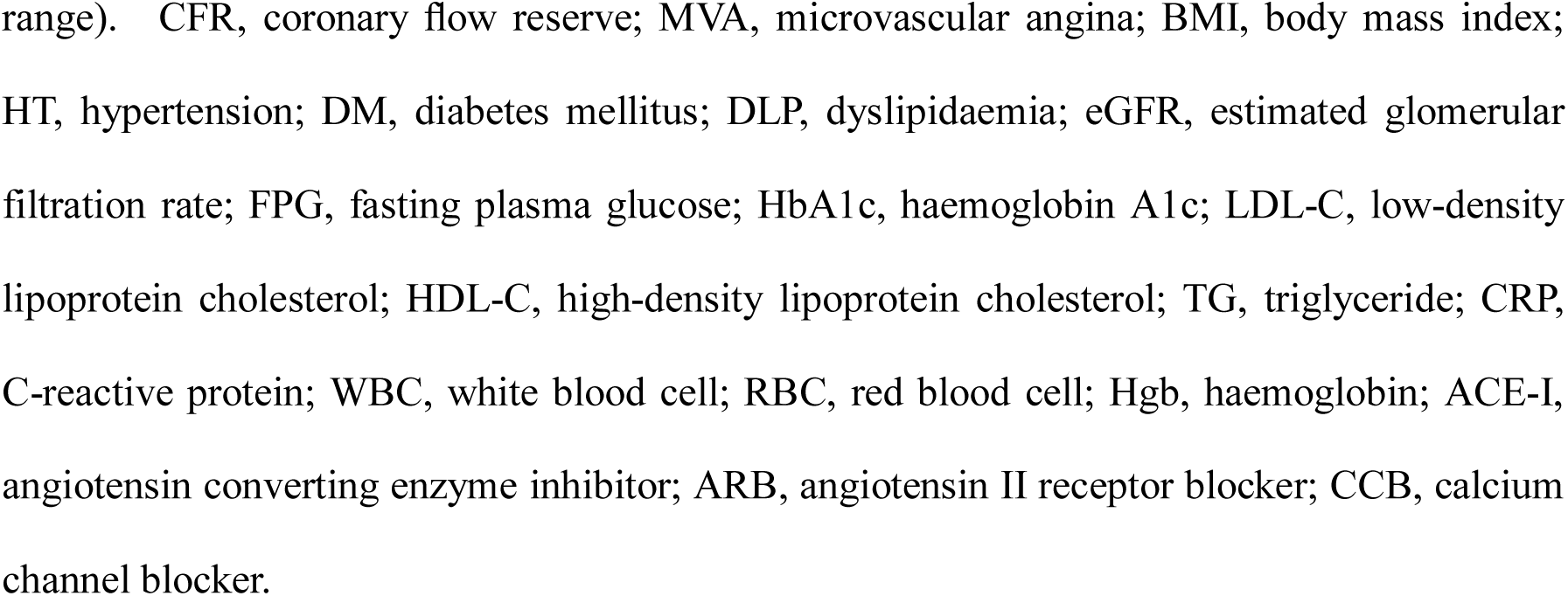
Clinical Characteristics of 55 MVA Patients Stratified into Those with a CFR <2.0 and Those with a CFR ≥2.0.

|  | **CFR <2.0**<br>(n=20) | **CFR ≥2.0**<br>(n =35) | p value |
| --- | --- | --- | --- |
| Age (years) | 64.6±12.9 | 62.3±11.8 | 0.52 |
| Sex (women), (%) | 9 (45%) | 29 (83%) | 0.006 |
| BMI (kg/m<sup>2</sup>) | 23.3±4.1 | 24.0±4.2 | 0.51 |
| Current smoking (%) | 1 (5%) | 6 (17%) | 0.40 |
| HT (%) | 7 (35%) | 20 (57%) | 0.16 |
| DM (%) | 3 (15%) | 7 (20%) | 0.73 |
| DLP (%) | 8 (40%) | 18 (51%) | 0.56 |
| eGFR (mL/min/1.73m<sup>2</sup>) | 71±15 | 73±15 | 0.72 |
| Uric acid (μmol/L) | 297±102 | 308±67 | 0.62 |
| FPG (mmol/L) | 5.6±1.1 | 5.3±1.2 | 0.47 |
| HbA1c (mmol/mol) | 42±8 | 44±12 | 0.46 |
| HbA1c excluding DM patients (mmol/mol) | 39±4 | 40±4 | 0.55 |
| LDL-C (mmol/L) | 2.8±0.7 | 3.0±0.8 | 0.49 |
| HDL-C (mmol/L) | 1.5±0.4 | 1.5±0.5 | 0.93 |
| TG (mmol/L) | 1.2 (0.8-1.6) | 1.2 (0.9-1.6) | 0.81 |
| CRP (mg/L) | 0.30 (0.28-1.33) | 0.80 (0.30-1.50) | 0.59 |
| WBC (×10<sup>9</sup>/L) | 6.1±2.2 | 5.4±1.2 | 0.11 |
| RBC (×10<sup>12</sup>/L) | 4.31 ±0.41 | 4.32±0.44 | 0.94 |
| Hgb (g/L) | 135±13 | 133±16 | 0.62 |
| Platelets (×10<sup>9</sup>/L) | 200±65 | 225±64 | 0.17 |
| Proteinuria (%) | 1/12 (8%) | 3/27 (11%) | 1.00 |
| <u>Medications (on admission)</u> |  |  |  |
| ACE-I or ARB (%) | 8 (40%) | 10 (29%) | 0.55 |
| CCB (%) | 10 (50%) | 24 (69%) | 0.25 |
| Nitrate (%) | 5 (25%) | 8 (23%) | 1.00 |
| Beta-blocker (%) | 6 (30%) | 6 (17%) | 0.32 |
| Statin (%) | 9 (45%) | 11 (31%) | 0.39 |

Data are number of patients (%), means ± standard deviation (SD), and medians (interquartile
1 range). CFR, coronary flow reserve; MVA, microvascular angina; BMI, body mass index; HT, hypertension; DM, diabetes mellitus; DLP, dyslipidaemia; eGFR, estimated glomerular filtration rate; FPG, fasting plasma glucose; HbA1c, haemoglobin A1c; LDL-C, low-density lipoprotein cholesterol; HDL-C, high-density lipoprotein cholesterol; TG, triglyceride; CRP, C-reactive protein; WBC, white blood cell; RBC, red blood cell; Hgb, haemoglobin; ACE-I, angiotensin converting enzyme inhibitor; ARB, angiotensin II receptor blocker; CCB, calcium channel blocker.

## Discussion

This case-control study investigated the clinical characteristics of patients with MVA. The major findings were as follows: (1) amongst 92 MVA patients, 39 were men (42%) and 53 were women (58%) (table 1 and figure 1); (2) elevated HbA1c, low HDL-C, and low Hgb levels were significantly associated with MVA in men and women (tables 4-5); (3) proteinuria was more common in men and women patients with MVA than in the control men and women, respectively (table 2); (4) MCV was significantly lower in men and women patients with MVA than in the control men and women, respectively (table 6); (5) in patients with MVA, a CFR ≥2.0 was more common in women than in men (table 7). To the best of our knowledge, this study is the first to report that elevated HbA1c, low HDL-C, and low Hgb levels were significantly associated with MVA, and a CFR ≥2.0 was more common in women with MVA than in men with MVA.

The mechanisms of CMD, which is the cause of MVA, are multifactorial and result from functional or structural abnormalities in the coronary microvasculature, including (1) increased vasoconstriction, (2) impaired coronary vasodilator capacities, and (3) increased coronary microvascular resistance, all three of which can occur alone or in combination ^1^. In this study, we enrolled 92 consecutive Japanese patients with MVA who underwent comprehensive evaluation of coronary microvascular function that consisted of cardiac catheterisation with an ACh provocation test and CFR measurements (interventional diagnostic procedure) to assess CMD. We evaluated the clinical characteristics of the MVA patients.

This study showed that an elevated HbA1c level was significantly associated with MVA. This result was true for both men and women (tables 3-5). The mean HbA1c levels in MVA patients were 44±11 mmol/mol, 43±11 mmol/mol, and 44±11 mmol/mol in men+women, men, and women, respectively, which are not severely hyperglycaemic levels (tables 1-2). Some reports have shown that diabetic microvascular diseases, such as retinopathy or nephropathy, can precede the appearance of type 2 diabetes, and may contribute to the development of micro-/macro-vascular complications in patients with type 2 DM, prediabetes, or hyperglycaemia ^17^ ^18^. Another study has shown that proteinuria in prediabetes might be an early biomarker for detectable diabetic nephropathy ^19^. This study found no differences of the prevalence rates of DM between the patients with MVA and the control participants. However, in this study, the number of proteinuria was significantly higher in patients with MVA than in the control participants. The HbA1c levels excluding DM patients were significantly higher in patients with MVA than in the control participants. (tables 1-2). These findings suggested that hyperglycaemia reflected by mildly elevated HbA1c levels such as in patients with prediabetes could be critically involved in the development of MVA. Hyperglycaemia may contribute to microvascular diseases by the suppression of endothelium-dependant vasodilation through the increased production of free radicals ^13^. Therefore, microvascular diseases associated with hyperglycaemia, as reflected by mildly elevated HbA1c levels (i.e., prediabetes), could represent systemic phenomena that extend beyond pathologic manifestations such as nephropathy to the coronary circulation. Mildly elevated HbA1c levels could be a critical marker for MVA.

This study showed that a low Hgb level was significantly associated with MVA. This result was true for both men and women (tables 3-5). A previous paper reported that reduced haemoglobin could predict adverse outcomes in patients with ischaemic heart disease not only through reduced oxygen delivery but also through reduced nitric oxide availability ^20^. In this study, although the Hgb levels did not satisfy the criterion for anaemia, Hgb and MCV levels were significantly lower in patients with MVA than in control participants, in both men and women (table 6). The serum iron level was significantly lower in the MVA women patients than in the control women. By contrast, the difference between the ferritin levels in MVA women patients and control women was not significant. Based on these observations, haematopoietic dysfunction caused by an iron disorder could be associated with MVA, although the aetiology could not be clarified by the available data in this study.

This study found that a low HDL-C level was significantly associated with MVA in both men and women (tables 4-5). Low HDL-C is an established risk factor not only for macrovascular events, but also for microvascular diseases in diabetic patients ^21^. The underlying mechanisms involved in the macrovascular and microvascular diseases in diabetes have been found to be the association of low HDL-C levels with increased oxidative stress, increased inflammation, and dysfunctional endothelium ^22^. Another study has shown that HDL-C could be a factor involved in reduced CFR ^8^. Based on these findings altogether, low HDL-C could be associated with MVA.

In this study, of 55 patients with MVA who underwent CFR measurement, a CFR ≥2.0 was more common in women than in men (29 women with CFR ≥2.0 [29/35 =83%] vs. 9 women with CFR <2.0 [9/20 =45%], p =0.006) (table 7). MVA patients with a CFR ≥2.0 had ischaemic findings during anginal symptoms and on ECG without epicardial coronary spasms by the ACh provocation test. The results suggested that these patients might have coronary microvascular spasms, although coronary microvascular spasms cannot be directly visualised. Our finding that coronary microvascular spasm was more common in women than in men was consistent with the findings of previous studies ^23^ ^24^. In our study, the women with MVA and a CFR ≥2.0 were aged 62.5 ±11.4 years (data not shown), which suggested that many of them were postmenopausal and had oestrogen deficiency. Oestrogen can modulate coronary microvascular vasomotor function ^13^. There are several types of MVA, which include increased vasoconstriction (microvascular spasm), impaired vasodilator capacity, and increased resistance ^1^. The different types of MVA might be sex specific. Further studies on the association of MVA subtypes with the sex of study patients are needed.

MVA can be detected in 30% to 50% of patients with anginal symptoms and nonobstructive CAD on invasive CAG ^5^ ^14^. CMD leading to MVA can play an important role not only in angina, but also in various cardiovascular diseases such as acute coronary syndrome and left ventricular hypertrophy ^13^ ^25^. Patients with anginal symptoms and angiographically nonobstructive CAD currently have large burdens of cardiovascular disease, including repeated hospitalisations and CAG, which also indicate that the economic burden of MVA is increasing ^26^. The majority of MVA patients remain undertreated because of the lack of evidence-based data. This study showed that hyperglycaemia as reflected by mildly elevated HbA1c levels, and low HDL-C and Hgb levels were significantly associated with MVA, and that a CFR ≥2.0 was more common in women than in men patients with MVA. Because there is currently no specific treatment for MVA, the management of modifiable risk factors, including conventional risk factors, can play an important role in the treatment of MVA ^4^ ^13^. Therefore, our observational findings may be helpful for further risk stratification and the development of treatment strategies for MVA.

### Study Limitations

This study has several limitations. First, it was a single-centre study with a relatively small sample size. Second, it was a retrospective study. Third, the study only included Japanese patients and participants. Fourth, our results may not be applicable to patients who are outside of the age range we studied. Fifth, the control group comprised 691 age-matched Japanese participants (466 men, 225 women) without a history of either chest pain or cardiovascular diseases. We did not have any information on the medications taken by the control participants. Moreover, we could not completely rule out that a control participant might have asymptomatic CAD. Sixth, we did not measure CFR in all the study patients and all the control participants. Further analyses and recalibrations of a larger and independent study population are needed.

## Conclusions

Hyperglycaemia reflected by mildly elevated HbA1c levels, (i.e., prediabetes) may be a risk factor for MVA. Low HDL-C and low Hgb levels may also be risk factors for MVA. CFR ≥2.0 was more common in women than in men amongst MVA patients. MVA could be a systemic phenomenon associated with microvascular diseases. The study findings may aid in identifying high-risk patients with MVA, allowing increased monitoring and treatments for them.

## Contributions

SS, KY, MI, HY, HO, and KT designed the study. SS, KY, MI, EM, NK, KH, NT, TA, KF, SH, KO, JS, and KT collected the data. SS, KY, MI, YA, HU, EY, KO, and JS carried out the statistical analyses and interpreted the data results. SS, KY, MI, and HY wrote the manuscript. EY, HS, HK, HJ, KK, HY, HO, and KT supervised the project. SS, KY, and MI contributed equally to this article. All authors read and approved the final manuscript.

## Patient and public involvement

Patients and/or the public were not involved in the design, or conduct, or reporting, or dissemination plans of this research.

## Patients consent for publication

Not applicable.

## Ethics approval

This study was conducted in accordance with the principles contained in the Declaration of Helsinki. The protocol of our study was approved by the ethics committee of the Faculty of Life Sciences, Kumamoto University, and the Japanese Red Cross Kumamoto Hospital Health Care Centre (approval number: 473). We obtained a signed informed consent from each patient and participant. This study is registered at the University Hospital Medical Information Network (UMIN) Clinical Trials Registry (UMIN 000043834). Opt-out materials are available through the website: https://kumadai-junnai.com/archives/clinical.

## Provenance and peer review

Not commissioned; externally peer reviewed.

## Data availability statement

The data that support the findings of this study are available upon reasonable request.

## Acknowledgement

We thank Michiyo Saito, MT, at our institution for her excellent technical assistance.

